# Effects of adiposity on the human plasma proteome: Observational and Mendelian randomization estimates

**DOI:** 10.1101/2020.06.01.20119081

**Authors:** Lucy J. Goudswaard, Joshua A. Bell, David A. Hughes, Laura J. Corbin, Klaudia Walter, George Davey Smith, Nicole Soranzo, John Danesh, Emanuele Di Angelantonio, Willem H. Ouwehand, Nicholas A. Watkins, David J. Roberts, Adam S. Butterworth, Ingeborg Hers, Nicholas J. Timpson

**Affiliations:** Medical Research Council (MRC) Integrative Epidemiology Unit at the University of Bristol, Bristol, UK; Population Health Sciences, Bristol Medical School, University of Bristol, Bristol, UK; School of Physiology, Pharmacology and Neuroscience, University of Bristol, UK; Wellcome Sanger Institute, Hinxton, UK; Department of Haematology, School of Clinical Medicine, University of Cambridge, UK; MRC/BHF Cardiovascular Epidemiology Unit, Department of Public Health and Primary Care, University of Cambridge, Cambridge, United Kingdom; NIHR Blood and Transplant Research Unit in Donor Health and Genomics, Department of Public Health and Primary Care, University of Cambridge, Cambridge, UK; British Heart Foundation Centre of Research Excellence, University of Cambridge, Cambridge, UK; NIHR Cambridge Biomedical Research Centre, University of Cambridge and Cambridge University Hospitals, Cambridge, UK; Health Data Research UK Cambridge, Wellcome Genome Campus and University of Cambridge, Cambridge, UK; NHS Blood and Transplant, Cambridge Biomedical Campus, Long Road, Cambridge, UK; NHS Blood and Transplant-Oxford Centre, Level 2, John Radcliffe Hospital, Oxford, UK; Radcliffe Department of Medicine, University of Oxford, John Radcliffe Hospital, Oxford, UK; Bristol Heart Institute, Bristol, UK

## Abstract

Variation in adiposity is associated with cardiometabolic disease outcomes, but the mechanisms leading from this exposure to disease are unclear. This study aimed to estimate effects of adiposity, proxied by body mass index (BMI), on 3,622 unique plasma proteins measured by the SomaLogic platform in 2,737 healthy participants from the INTERVAL study of UK blood donors. We conducted both observational and Mendelian randomization analyses where we used a genetic risk score for BMI as an instrument to estimate effects of BMI on protein levels. Our results suggest that BMI has a broad impact on the human plasma proteome, with estimated effects of BMI appearing strongest on proteins including circulating leptin, sex hormone-binding globulin and fatty acid-binding protein-4. We also provide evidence that proteins most altered by BMI are enriched for genes involved in cardiovascular disease. Altogether, these results help to focus attention onto new potential proteomic signatures of obesity-related disease.

## Introduction

Obesity has tripled worldwide since 1975, now affecting around 40% of adults in the United States and 26% of adults in the UK (2). The average body mass index (BMI) of the UK adult population is now in the conventional ‘overweight’ category (BMI between 25 and 30 kg/m^2^) (3) and ‘overweight’ is now more common than ‘normal-weight’ in middle age in many high-income countries (4). BMI is often used as a proxy for adiposity given high correlations between BMI and more objectively measured fat mass indices (5). Higher adiposity is a major risk factor for various noncommunicable diseases including type II diabetes, cardiovascular diseases, musculoskeletal diseases, and cancer (6-9), which collectively put a substantial strain on health services (10, 11). These BMI-disease associations are supported by prospective observational studies and, more recently, by Mendelian randomization (MR) studies (12-14), which use the naturally random assignment of adiposity-associated risk alleles to estimate effects of BMI on disease outcomes. This method helps to overcome issues such as confounding and reverse causation which commonly occur with observational studies (15).

Despite MR studies supporting a causal role of adiposity for cardiometabolic diseases, and randomised trials supporting the effectiveness of weight loss in reducing disease risk (16), the molecular footprint of adiposity is not well understood. Previous studies have largely focused on the impact of higher BMI on the lipidome including traits such as cholesterol and triglycerides in lipoprotein subtypes (e.g. low-density lipoprotein (LDL), very-low-density lipoprotein (VLDL), and high-density lipoprotein (HDL) particles) (5, 17), and on inflammatory molecules such as C-reactive protein (CRP) (17, 18). Efforts to study the effect of BMI on the broader proteome have generally been in an observational framework (19), with no studies using MR to investigate the impact of BMI on a comprehensive set of protein traits measured in circulation. This has only recently become possible with newly developed proteomic technologies such as the SomaLogic platform. It is estimated that around 25% of proteins in the human proteome circulate in blood (20), which allows SomaLogic to quantify circulating traits including enzymes, protein kinases and transport proteins in unprecedented scope and detail (21). This research is important because the majority of druggable targets are such proteins (22). Identifying proteins which are altered by BMI may allow for the development or repurposing of therapeutic interventions to reduce the impact of BMI on disease.

In this study, we aimed to measure associations between adiposity and the human proteome and to also estimate the underlying effects in a causal framework. Using data on 2,737 participants from a cohort study of blood donors in England (INTERVAL), we estimated effects of BMI on 4,034 (3,622 unique) plasma protein traits, in both observational and MR frameworks. We examined the agreement between effect estimates from different methods and then performed enrichment analyses of the most strongly altered proteins to map their potential relevance to cardiovascular disease.

## Methods

### Study population

INTERVAL is a prospective cohort study which was initially a randomised trial that aimed to test the efficiency and safety of reducing the time between whole blood donation in approximately 50,000 participants (1). Upon informed consent, eligible participants who were: aged 18 years and over, willing to complete online questionnaires and without a self-reported history of major disease were recruited between June 11^th^, 2012 and June 15^th^, 2014 from 25 National Health Service Blood and Transplant (NHSBT) centres across England. Participants filled out questionnaires including self-reported height and weight, smoking frequency and alcohol consumption. Blood samples were taken at baseline which were analysed for full blood counts and blood biomarkers. This study was approved by Cambridge (East) Research Ethics Committee. Access to the data was granted by the Data Access Committee. Data cannot be publicly shared due to sensitive content. General enquiries can be sent to the INTERVAL team https://www.intervalstudy.org.uk/more-information/.

The present study was conducted on a random subset of participants from INTERVAL who had plasma proteins measured by SomaLogic. This included up to 2,737 participants mostly of European descent across analyses described below.

### Assessment of BMI and covariables

Participants completed online questionnaires wherein they reported their height and weight. BMI was calculated as weight in kilograms divided by the square of their height in metres (kg/m^2^). Available covariables were age, sex, previous or current smoking frequency (in three categories of: never, occasional, most days or every day) and alcohol intake frequency (in four categories of: rarely, less than once a week, 1-2 times a week, 3-5 times a week or most days). These covariables were chosen as they were measured in the INTERVAL collection and are measures which have known relationships with adiposity and cardiometabolic health.

### Measurement of circulating proteins

Plasma proteins were measured in INTERVAL participants at baseline (before randomisation of assignment to the time interval between blood donation) using the SomaScan^®^ by SomaLogic. This platform uses 4,034 modified nucleotides known as Slow Off-rate Modified Aptamers (SOMAmers) which make direct contact with proteins, enabling detection of 3,622 unique proteins or protein complexes and quantifies them using a DNA microarray (21). These proteins include growth factors, cytokines, protein kinases, and transport proteins. The proteins were measured in relative fluorescence units (RFUs) and quality control (QC) was performed as described by Sun et al. (23).

There was no missingness across protein variables. The proteomic data used had been pre-adjusted for: age, sex, duration between blood draw and sample processing (1 day or less vs >1 day), and the first three genetic principal components, with the residuals inverse normal rank transformed.

### Genetic data and instrument for BMI

INTERVAL participant genotyping was performed on the Affymetrix GeneTitan® Multi-Channel (MC) Instrument and the QC of genotype data was implemented as described by Astle et al. (24). The imputation panel used was the 1000 genomes phase-3-UK-10K (24). A genetic instrument for BMI was constructed using 654 genetic variants that were associated with BMI at P < 5 x 10^−8^ in the inverse variance weighted fixed-effect meta-analysis of GWAS of ∼700,000 individuals of European ancestry (25). This meta-analysis consisted of ∼250,000 adults from the Genetic Investigation of ANthromopetric Traits (GIANT) consortium (26) and ∼450,000 adults from the UK Biobank study. Only 0.05% of UK Biobank participants were included in the INTERVAL study. These participants were not excluded to increase power. The weighted GRS was made using PLINK 2.0 software (27) using the effect alleles and beta coefficients from the source GWAS. The score was calculated by multiplying the number of effect alleles at each SNP by its weighting or effect estimate (beta), summing these, and dividing by the total number of SNPs included. The GRS therefore can be interpreted as the average per-SNP effect on BMI for each individual.

### Statistical analyses

The population characteristics of INTERVAL participants with SomaLogic data who were included in this study (N range: 2,422 to 2,737) were compared to those INTERVAL participants who were not included (N range: 27,174 to 30,721). Population characteristics evaluated were age, sex, weight, height, BMI, smoking frequency, and alcohol intake. Differences among the two INTERVAL sub-sets were tested by a two-sided t-test for continuous traits and a two-sided Chi-square test for categorical variables. Observational analyses were conducted using linear regression models to examine associations between BMI (in normalized standard deviation (SD) units based on a rank normal transformation) and each standardised protein trait as a dependent (outcome) variable. Two models were used: 1) adjusted for age and sex and 2) additionally adjusted for smoking and alcohol consumption (each as an ordered categorical variable). These estimates therefore reflect the normalized SD-unit difference in each protein trait per normalized SD-unit (4.8 kg/m^2^) higher BMI. Associations of covariables with BMI and protein traits were also examined using linear regression to affirm their potential roles as confounders of associations between BMI and protein traits.

A Shapiro-Wilk test was used to test whether the GRS showed a normal distribution. MR analyses were conducted using two-stage least squares (2SLS) regression models with robust standard errors, with measured BMI in SD units and the GRS for BMI as the instrumental variable. These MR estimates reflect the normalized SD-unit difference in each protein trait per normalized SD-unit (4.8 kg/m^2^) higher BMI. Agreement between observational and MR estimates was examined using a separate linear regression model (based on the beta coefficients from observational and MR analyses), with a regression slope nearer one and a higher R^2^ reflecting stronger agreement between estimates. This was performed: 1) for all proteins, and 2) excluding the proteins that fell below our P-value reference point for strong evidence (defined below) to examine whether agreement is limited to ‘top hits’ or applies throughout the effect distribution.

Given that multiple statistical tests were performed, a Bonferroni correction was used to penalise results. This was informed by the correlation between proteins, adjusting only for the estimated number of independent traits (**Supplementary Fig 1**). Correlation was assessed by a Spearman’s correlation matrix. From a starting number of 4,034, the number of independent proteins was 3,655 (using a correlation cut-off of r = 0.8 / tree cut height = 0.2 between proteins, **Supplementary Fig 2**). As a heuristic for grading the strength of evidence given current sample sizes, we utilise a Bonferroni adjusted P-value of < 1.4 x 10^5^ (which corresponds to an approximate 0.05 level (0.05/3655)) to indicate strong evidence in this sample. Results are presented in full in the supplementary material with exact P-values, effect sizes, and 95% confidence intervals.

### Enrichment analysis

To investigate whether any proteins showed a particularly strong relationship with BMI and disease (signal detection), protein features were clustered for further analysis. Firstly, a principal component analysis (PCA) was performed on the inverse normal rank transformed protein dataset to generate components explaining correlated features and to reduce the dataset. The number of principal components (PCs) was chosen to convey most of the dataset while reducing the number of variables; this was selected by visualising the ‘elbow’ of a scree plot (**Supplementary Fig 3)**. Derived PCs were then entered into a k-means analysis to create clusters of similar proteins. The number of clusters was chosen as the ‘elbow’ of a scree plot of the number of protein groups against how much variance of the dataset is explained by clusters.

To explore whether there was a systematic difference in the association of proteins within these clusters and BMI, the beta coefficients from the observational linear regressions or MR models were transformed into their absolute values and divided by their standard error (SE). The mean of the absolute beta divided by SE in each cluster was compared using a one-tailed pairwise Wilcox test to identify which clusters showed a stronger association with BMI. For the cluster(s) showing evidence for larger absolute beta coefficients, an enrichment analysis was performed using DAVID bioinformatics resources 6.8 (28). Enrichment was assessed by using the uniprot IDs for the proteins in the cluster and comparing these proteins with the uniprot IDs of the full SomaLogic protein list. Enrichment for protein involvement in disease (using the genetic association database (GAD) disease classes (29)) of the protein cluster was assessed by fold enrichment and a Bonferroni-corrected P value to account for multiple testing. As well as looking at cluster enrichment, the proteins that were associated with BMI in confounder-adjusted observational analyses at P < 1.4 x 10^−5^ were entered into the disease enrichment tool and compared with the total proteins (as described for cluster enrichment).

Analyses were performed using R version 3.4.2 (30).

## Results

### Participant characteristics

INTERVAL participants included in this study (those with proteomic data), had a mean age of 45.0 years (SD of 14.1 years) and 48.3% were female (**Table 1**). Mean BMI was 25.9 kg/m^2^ (SD of 4.8 kg/m^2^) and the majority of participants were non-smokers (59.1%). Nearly a quarter (23.5%) reported currently or previously smoking daily and 71.5% reported drinking alcohol at least once a week.

**Table 1.**
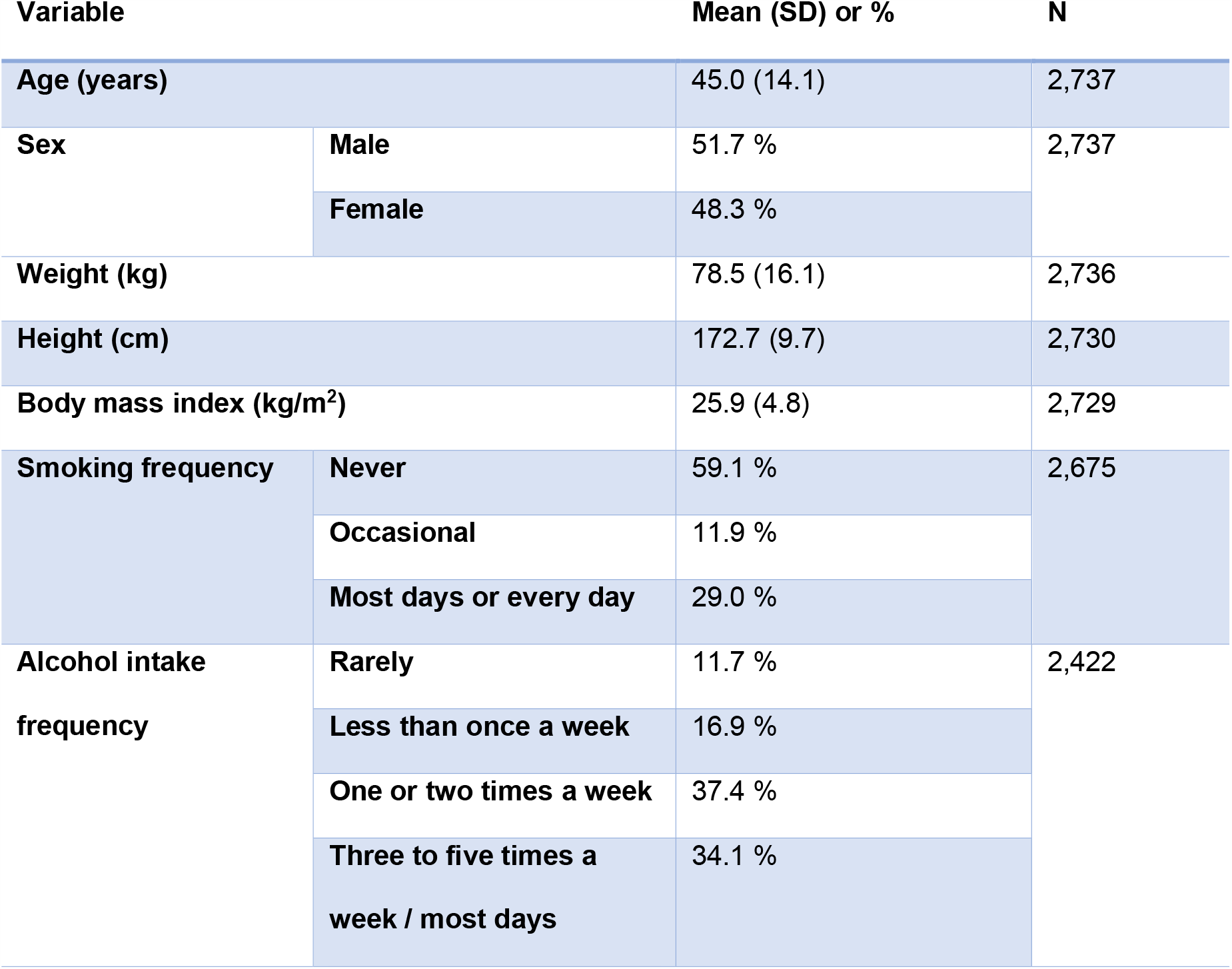
Characteristics of included participants. Compared with participants who were included in analyses, those who were excluded were of similar age, had a similar BMI and similar drinking and smoking habits (**Supplementary Table 1**). There was a slightly smaller proportion of females included in the analysis compared with excluded participants (48.3% and 50.3% respectively, P = 0.04).

| Variable |  | Mean (SD) or % | N |
| --- | --- | --- | --- |
| Age (years) |  | 45.0 (14.1) | 2,737 |
| Sex | Male | 51.7 % | 2,737 |
|  | Female | 48.3 % |  |
| Weight (kg) |  | 78.5 (16.1) | 2,736 |
| Height (cm) |  | 172.7 (9.7) | 2,730 |
| Body mass index (kg/m<sup>2</sup>) |  | 25.9 (4.8) | 2,729 |
| Smoking frequency | Never | 59.1 % | 2,675 |
|  | Occasional | 11.9 % |  |
|  | Most days or every day | 29.0 % |  |
| Alcohol intake frequency | Rarely | 11.7 % | 2,422 |
|  | Less than once a week | 16.9 % |  |
|  | One or two times a week | 37.4 % |  |
|  | Three to five times a week / most days | 34.1 % |  |

### Observational estimates of associations of BMI with protein traits

In a linear regression model adjusted for age and sex among 2,729 adults, BMI (per SD higher) was associated with 1,576 proteins (39%) at the level P < 1.4 x 10^−5^ (multiple testing reference point) **Supplementary Table 2**). In a second model additionally adjusting for frequencies of smoking and alcohol intake among 2,380 adults, there were 1,447 associations at the same reference point (**Supplementary Table 3**). The strongest positive associations were with leptin (0.74 SD, 95% CI 0.71 to 0.77, P = 9.9 x 10^−324^) and adipocyte fatty acid binding protein (FABP4) (0.58 SD, 95% CI 0.55 to 0.62, P = 6.4 x 10^−211^). BMI (per SD) was also strongly positively associated with inflammatory proteins such as Complement Factor I (0.46 SD, 95% CI 0.43 to 0.50, P = 5.6 x 10^−122^) and CRP (0.44 SD, 95% CI 0.41 to 0.48, P = 8.2 x 10^−112^). BMI (per SD) also showed strong negative associations with proteins such as insulin-like growth factor-binding protein (IGFBP) 2 (−0.48 SD, 95% CI −0.51 to − 0.44, P = 2.7 x 10^−133^) and sex hormone-binding globulin (SHBG) (−0.43 SD, 95% CI −0.47 to −0.39, P = 2.4 x 10^−106^).

### Observational associations of covariables with BMI and protein traits

Age, sex, and frequencies of smoking and alcohol intake were each associated with BMI (**Supplementary Table 4**). Males had a higher BMI than females (0.17 SD, 95% CI 0.10 to 0.25, P = 5.8 x 10^−6^). Age was positively associated with BMI (0.01 SD higher per year older, 95% CI 0.009 to 0.015, P = 1.2 x 10^−18^). Smoking frequency was also positively associated with BMI (0.09 SD higher BMI per increase in smoking category, 95% CI 0.05 to 0.13, P = 3.2 x 10^−5^). Alcohol intake frequency was negatively associated with BMI (−0.08 SD decrease in BMI per increase in alcohol intake category, 95% CI −0.12 to −0.04, P = 7.1 x 10^−5^). The exposures age, sex, and frequencies of smoking and alcohol intake each also showed associations with protein traits (**Supplementary Tables 5-8** and **Supplementary Figs 4A-D)**.

### Associations of the GRS for BMI with measured BMI and covariables

The distribution of the GRS among participants was normal (mean = 0.08, SD = 0.29, W = 0.99, P = 0.73, **Fig 1A**). The GRS was associated with BMI in INTERVAL (based on self-reported height and weight), explaining 2.8% of its variance (R^2^ = 0.028, P = 1.6 x 10^−18^, **Fig 1B** and **Table 2**). There was no strong evidence of association between GRS and age (R^2^ = 0.001, P = 0.11), sex (R^2^ = 6 x 10^−5^, P = 0.28), smoking frequency (R^2^ = < 0.0001, P = 0.91), or alcohol intake (R^2^ < 0.0001, P = 0.44).

**Table 2.** Associations of the genetic risk score for BMI with reported BMI and covariables.

| Variable | N | Beta coefficient<br>(per 1-unit increase in GRS) | Standard error | P value | Adjusted $R^2$ | F statistic |
| --- | --- | --- | --- | --- | --- | --- |
| BMI (SDs) | 2729 | 0.57 | 0.06 | $1.64 \times 10^{-18}$ | 0.028 | 78.20 |
| BMI (kg/m<sup>2</sup>) | 2729 | 2.54 | 0.31 | $4.82 \times 10^{-16}$ | 0.024 | 66.68 |
| Age (years) | 2737 | -1.47 | 0.92 | 0.11 | 0.001 | 2.55 |
| Sex<br>(1 = female, 2 = male) | 2737 | -0.04 | 0.03 | 0.28 | $5.96 \times 10^{-5}$ | 1.16 |
| Smoking frequency | 2675 | -0.02 | 0.14 | 0.91 | -0.0004 | 0.01 |

| (1 = never, 2 = occasional, 3 = most days or every day) |  |  |  |  |  |  |
| --- | --- | --- | --- | --- | --- | --- |
| Alcohol intake frequency<br>(1 = rarely, 2 = < 1 a week, 3 = 1-2 times a week, 4 = most days or every day) | 2422 | -0.06 | 0.07 | 0.44 | -0.0002 | 0.60 |

**Fig 1.**
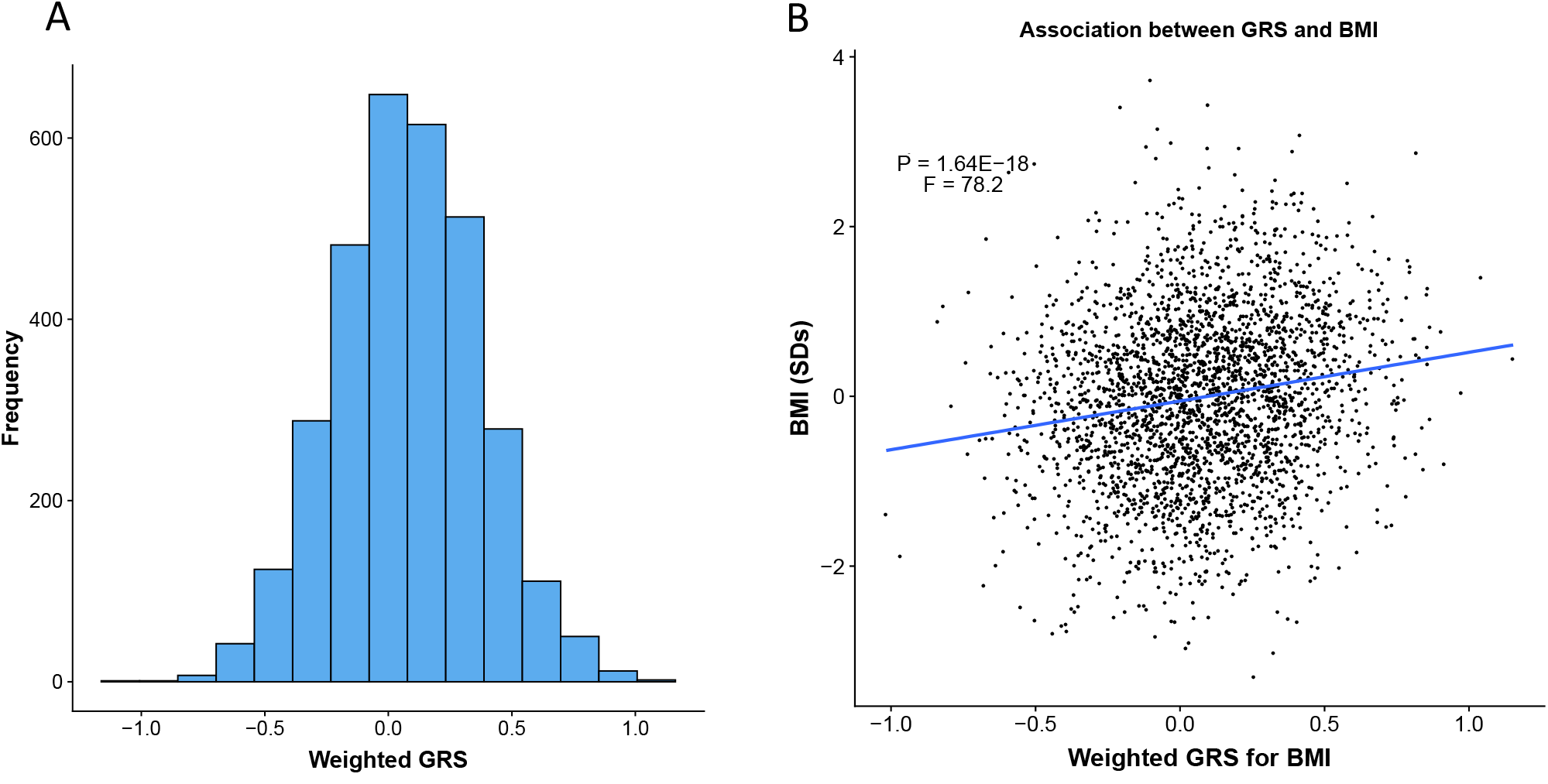
The genetic risk score shows a normal distribution and associates with reported BMI. **A**) Distribution of the genetic risk score for BMI in INTERVAL, **B**) Scatter plot of the genetic risk score for BMI against reported BMI in INTERVAL with the linear regression line (blue, N=2729).

### MR estimates of associations between BMI and protein traits

In MR analyses, eight unique BMI-protein associations were detected at the level P < 1.4 x 10^−5^ (multiple testing reference point, **Fig 2**). The strongest association of BMI (per SD) was again with leptin (0.63 SD, 95% CI = 0.48 to 0.79; P = 1.6 x 10^−15^); this was followed by the association with FABP4 (0.65 SD, 95% CI = 0.46 to 0.83; P = 6.7 x 10^−12^). A strong negative association was also seen between BMI (per SD) and SHBG (−0.45 SD, 95% CI −0.65 to −0.25, P = 1.4 x 10^−5^). Other BMI-protein associations (below a P-value reference point of 1.4 x 10^−5^) included positive associations with fumarylacetoacetase, inhibin β C chain and complement C5, and negative associations with receptor-type tyrosine-protein phosphatase delta and PILR alpha-associated neural protein. **Supplementary Table 9** provides the full MR results for BMI and protein traits.

**Fig 2.**
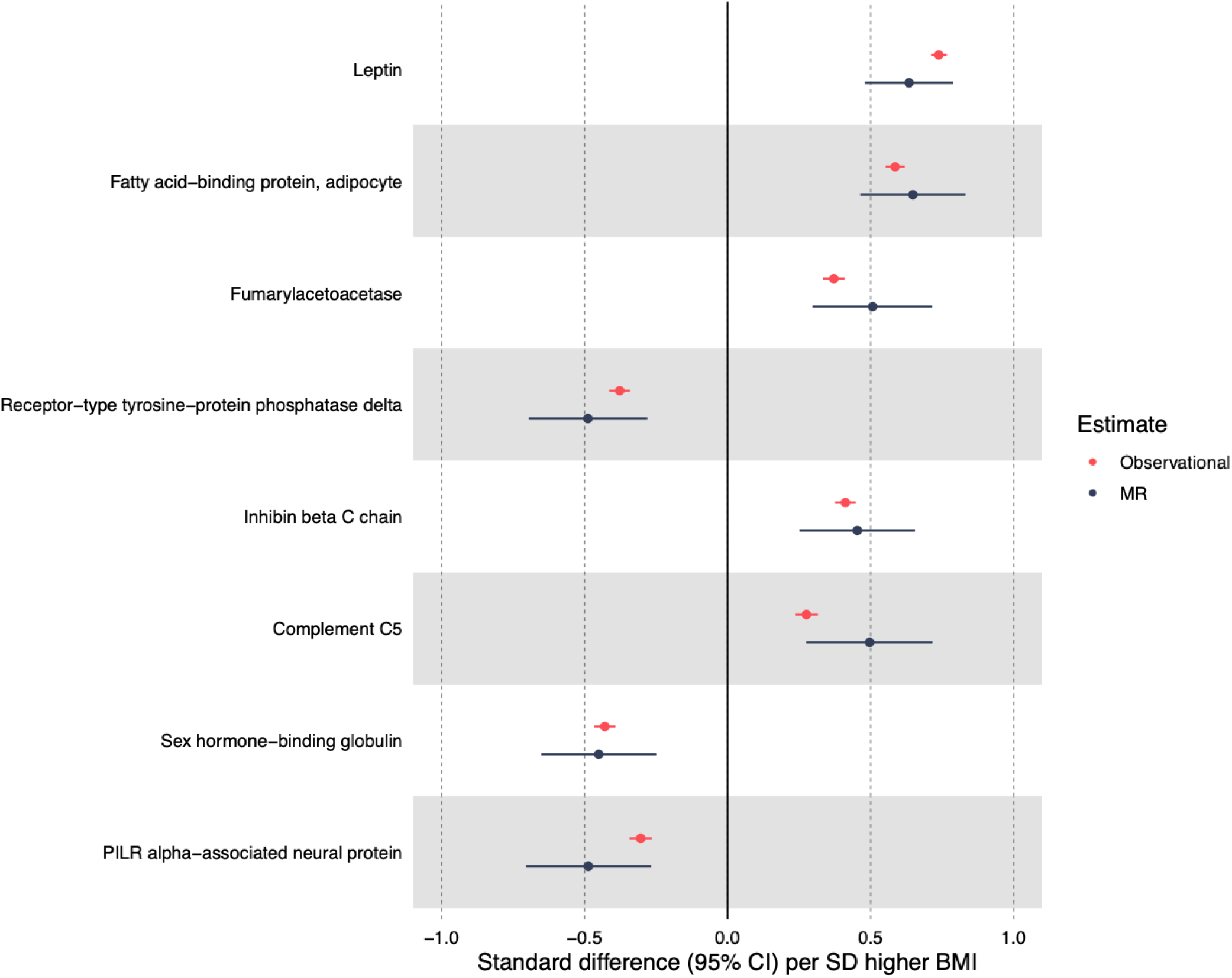
Strongest BMI and MR associations with corresponding observational associations. Strongest MR results of BMI and protein traits based on P value < 1.4 x 10^−5^ and their corresponding observational estimates.

### Comparison of observational and MR estimates

The distribution of P-values for associations between BMI and protein traits suggested an overrepresentation of signal for the observational estimates of BMI and protein traits; far more than expected from chance alone (**Supplementary Fig 5A**). In contrast to this, whilst there was still more signal than expected by chance for a large number of traits in MR analyses, the extent of this overrepresentation was reduced considerably (**Supplementary Fig 5B**).

The unadjusted and confounder-adjusted regression coefficients for BMI and protein traits were strongly associated (β = 0.99 SDs, R^2^ = 0.99, P = 9.9 x 10^−324^ **Fig 3A**). Compared with the observational estimates, the MR estimates were less precise, but there was a strong positive association between the beta coefficients from observational and MR estimates (β = 0.68 SDs, R^2^ = 0.33, P = 9.9 x 10^−324^) (**Fig 3B)**. After removing the proteins that were below the Bonferroni-corrected reference P-value of 1.4 x 10^−5^, the strength of association between unadjusted and adjusted observational estimates remained, but the association between observational and MR estimates attenuated slightly (**Supplementary Figs 6A/B**)

**Fig 3.**
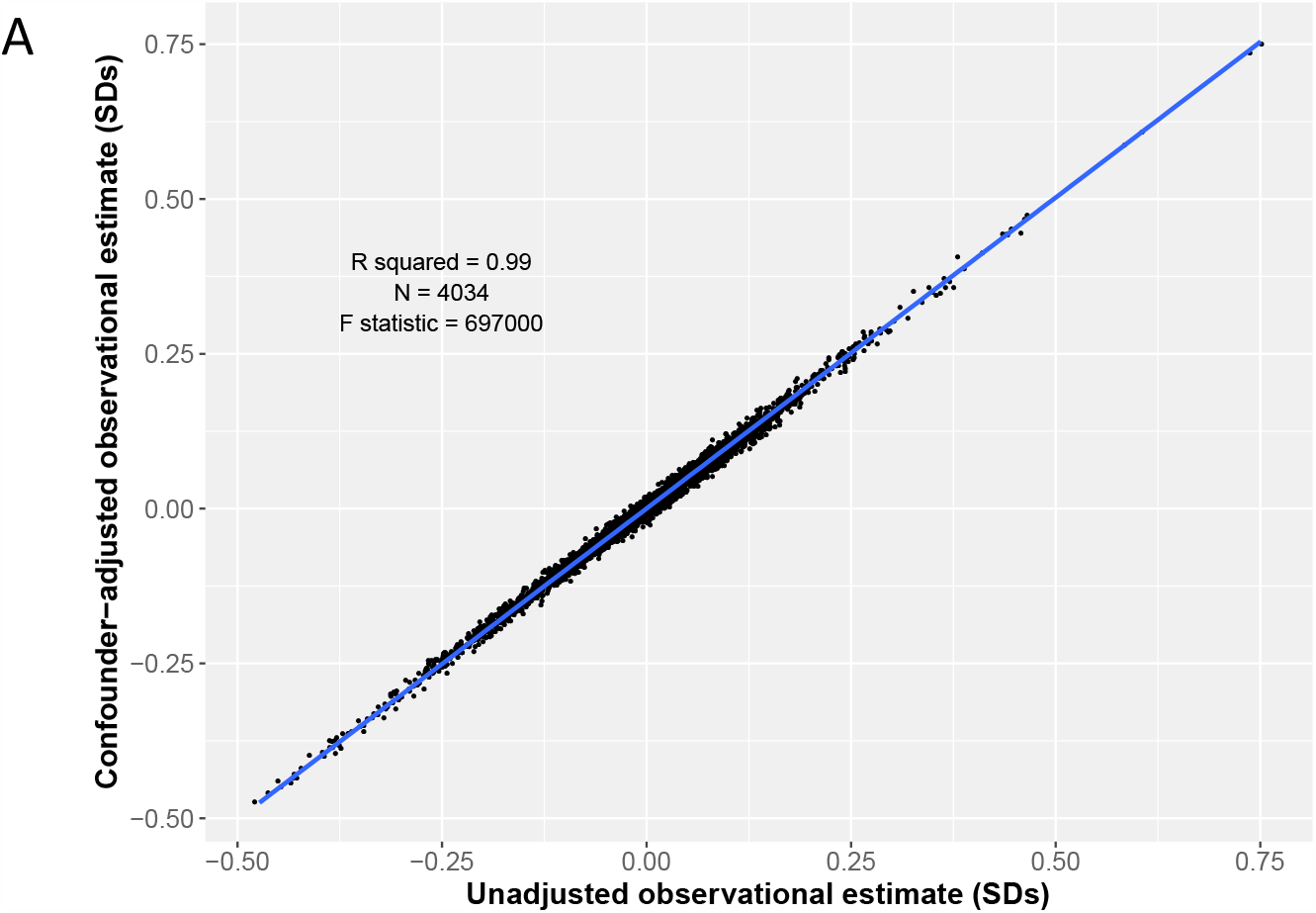
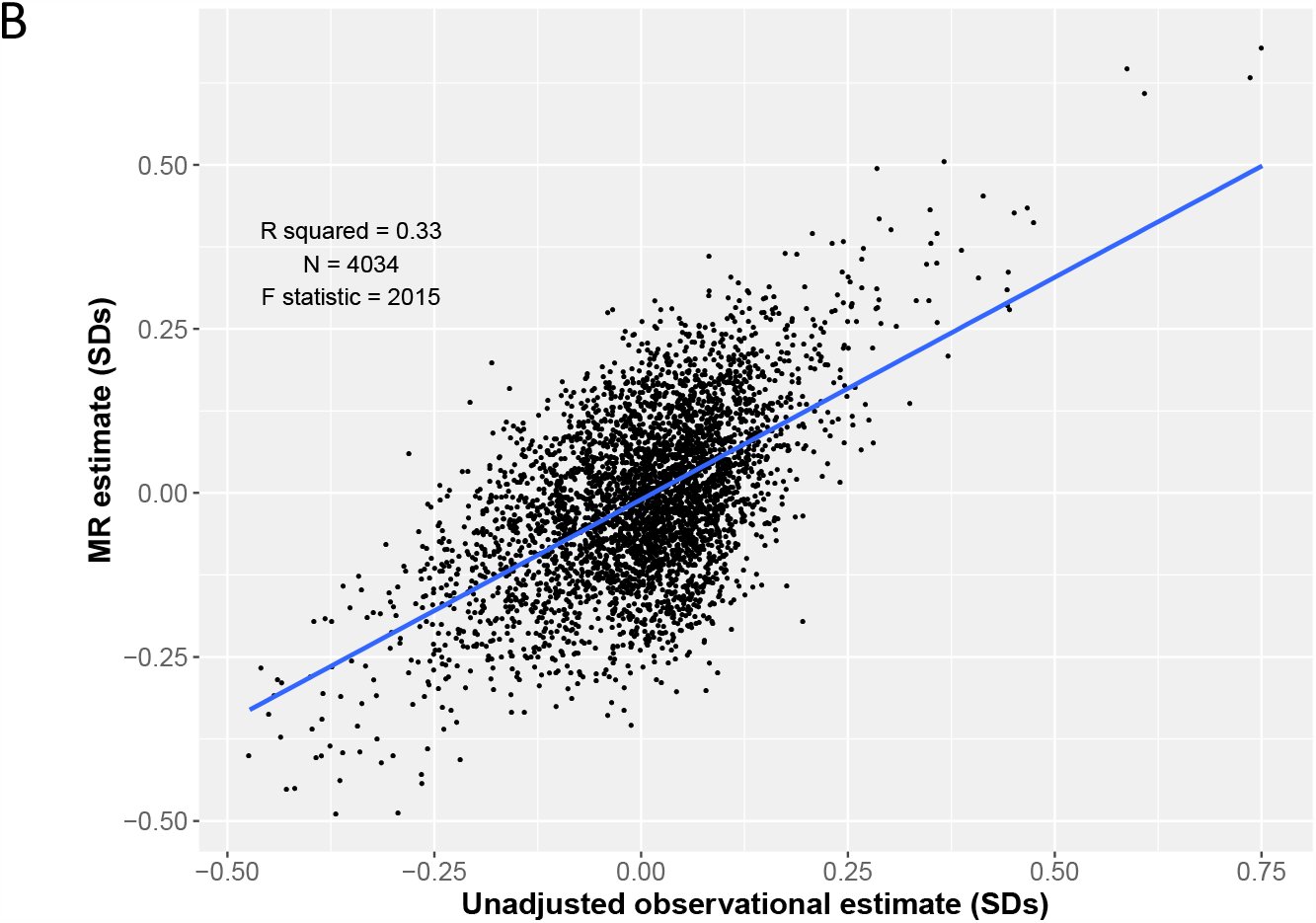
Observational and MR estimates show a positive association. **A**) Scatter plot of the unadjusted (age and sex adjusted) observational estimates and the confounder-adjusted observational estimates for BMI and protein traits with a regression line (blue). **3B**) Scatter plot of the unadjusted (age and sex adjusted) observational estimates and the MR estimates for BMI and protein traits with a regression line (blue).

### Enrichment analysis of strongest BMI-protein associations

In examining the clustering of proteins, visual representation using a scree plot suggested there were five PCs (**Supplementary Fig 3**). These five PCs were entered into a k-means analysis, which provided evidence for five clusters (grouping of individual proteins is included in **Supplementary Table 9**). To identify which cluster was most strongly affected by BMI, the mean absolute beta coefficient divided by the SE for each cluster was compared with the overall estimate. The beta coefficients for each BMI-protein effect, grouped by cluster can be seen in **Figs 4A** and **4B**. Six of the proteins out of the eight strongest BMI-protein MR estimates were in cluster 2 (**Supplementary Fig 9**). There was consistent evidence that cluster 2 showed a stronger association with BMI than the overall average BMI-protein effect both observationally (5.2 (SD 5) vs 4.38 (SD 4.16) respectively, P = 3.7 x 10^−4^) and in MR (1.04 (SD 0.84) vs 0.9 (SD 0.74), P = 5.3 x 10^−6^, **Supplementary Table 10, Figs 4C and 4D**). Cluster 2 showed consistent evidence of a having the largest BMI effect. Compared with the full protein list in SomaLogic, the proteins in cluster 2 were enriched for disease (**Table 3**), including cardiovascular disease (1.14 fold enrichment, P = 1.3 x 10^−4^), renal disease (1.22 fold enrichment, P = 1.0 x 10^−3^) cancer (1.1 fold enrichment, P = 9.5 x 10^−3^) and metabolic disease (1.08 fold enrichment, P = 4.2 x 10^−2^). No other individual cluster showed enrichment for disease. Enrichment for disease was also explored by comparing the proteins which had an association with BMI (P < 1.4 x 10^−5^) in the confounder-adjusted regression model with the total protein list. Compared with the full protein list, the proteins which showed a stronger observational association with BMI were enriched for renal disease (1.21 fold enrichment, P = 0.001) and metabolic disease (1.9 fold enrichment, P = 0.015, **Supplementary Table 11**).

**Table 3.**
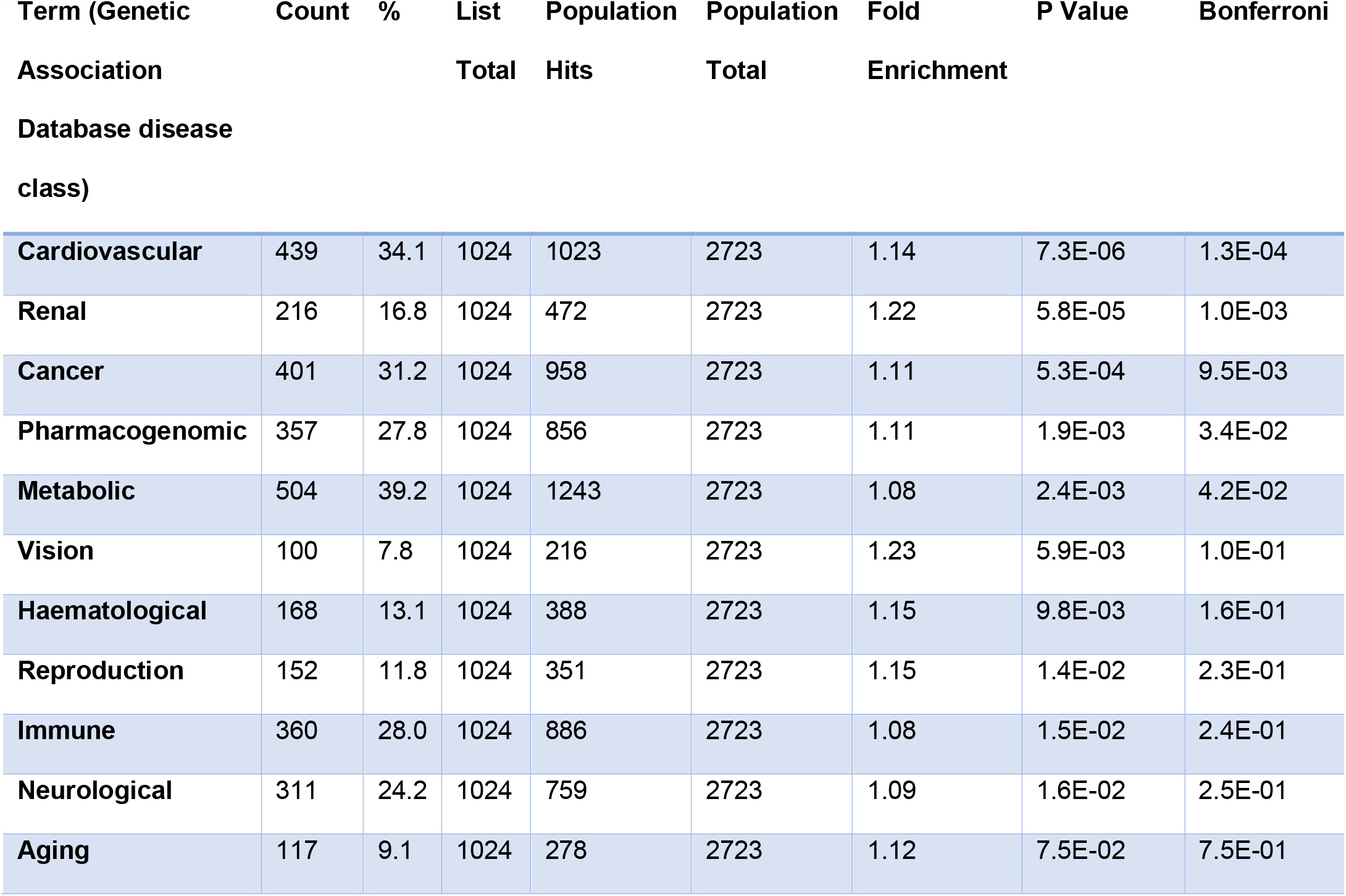
Cluster 2 vs full SomaLogic protein enrichment results for disease class using DAVID bioinformatics 6.8 (28)

| Term (Genetic Association Database disease class) | Count | % | List Total | Population Hits | Population Total | Fold Enrichment | P Value | Bonferroni |
| --- | --- | --- | --- | --- | --- | --- | --- | --- |
| Cardiovascular | 439 | 34.1 | 1024 | 1023 | 2723 | 1.14 | 7.3E-06 | 1.3E-04 |
| Renal | 216 | 16.8 | 1024 | 472 | 2723 | 1.22 | 5.8E-05 | 1.0E-03 |
| Cancer | 401 | 31.2 | 1024 | 958 | 2723 | 1.11 | 5.3E-04 | 9.5E-03 |
| Pharmacogenomic | 357 | 27.8 | 1024 | 856 | 2723 | 1.11 | 1.9E-03 | 3.4E-02 |
| Metabolic | 504 | 39.2 | 1024 | 1243 | 2723 | 1.08 | 2.4E-03 | 4.2E-02 |
| Vision | 100 | 7.8 | 1024 | 216 | 2723 | 1.23 | 5.9E-03 | 1.0E-01 |
| Haematological | 168 | 13.1 | 1024 | 388 | 2723 | 1.15 | 9.8E-03 | 1.6E-01 |
| Reproduction | 152 | 11.8 | 1024 | 351 | 2723 | 1.15 | 1.4E-02 | 2.3E-01 |
| Immune | 360 | 28.0 | 1024 | 886 | 2723 | 1.08 | 1.5E-02 | 2.4E-01 |
| Neurological | 311 | 24.2 | 1024 | 759 | 2723 | 1.09 | 1.6E-02 | 2.5E-01 |
| Aging | 117 | 9.1 | 1024 | 278 | 2723 | 1.12 | 7.5E-02 | 7.5E-01 |

**Fig 4.**
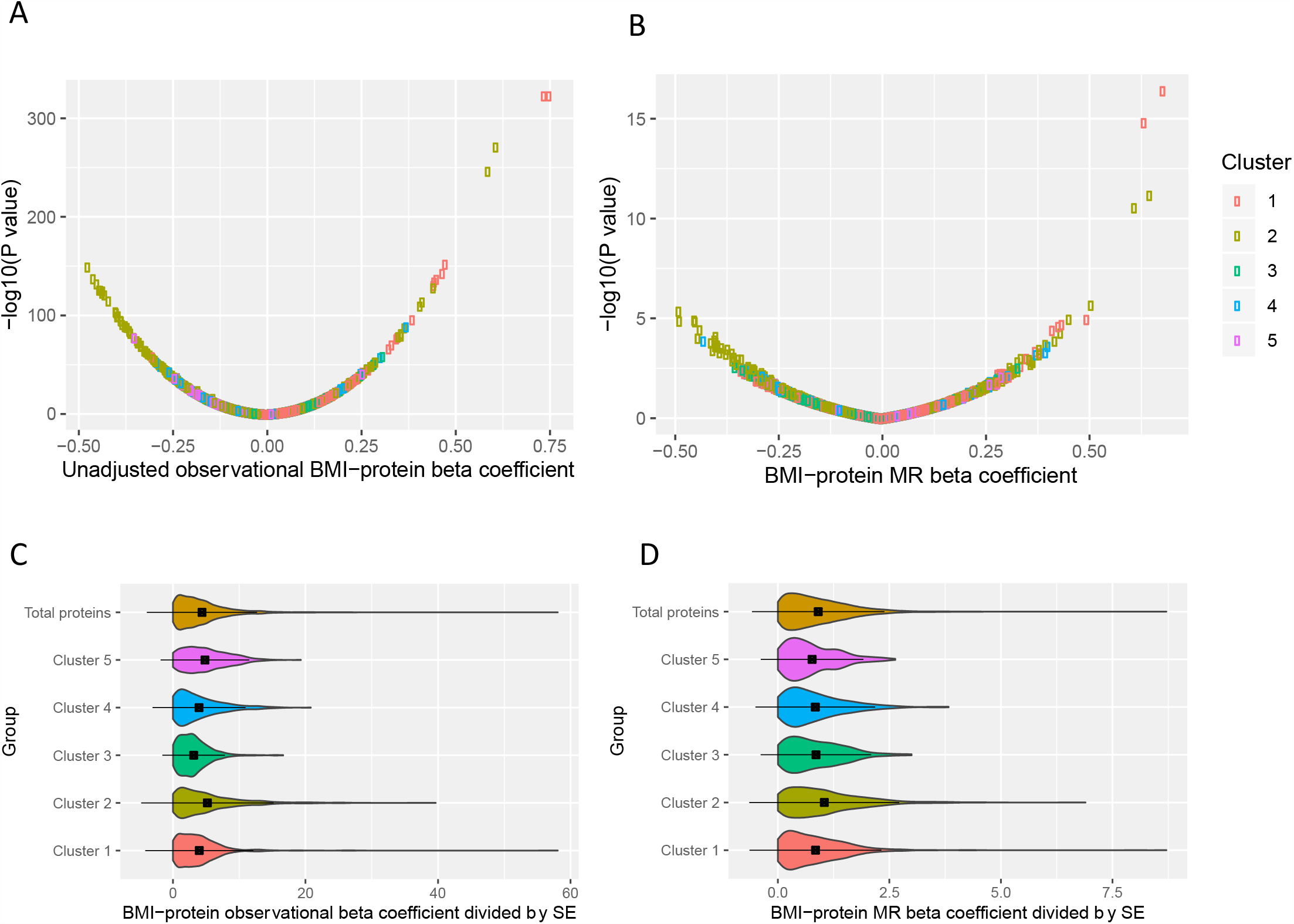
Cluster 2 shows larger absolute observational and MR BMI-protein estimates. **A)** Volcano plot showing the beta coefficients and -log_10_(P value) for each unadjusted observational BMI-protein association, with colour indicating the grouping by cluster. **B)** Volcano plot showing the beta coefficients and -log_10_(P value) for each BMI-protein Mendelian randomization (MR) estimate with colour indicating the grouping by clustering. **C)** Violin plot of the absolute beta coefficients divided by standard error for each BMI-protein association for the unadjusted observational model, grouped by cluster (mean ± SD plotted). **D)** Violin plot of the absolute beta coefficients for each BMI-protein Mendelian randomization estimate divided by the standard error, grouped by cluster (mean ± SD).

## Discussion

We aimed in this study to estimate the effects of adiposity on a comprehensive set of protein traits only recently measurable by untargeted proteomics using observational and MR methods. Our results suggest that adiposity alters numerous protein traits involved in regulating appetite, sex hormones, inflammation, and other systems; specific proteins most altered by BMI include leptin, FABP4, and SHBG. Results of follow-up analyses suggest that the cluster of proteins most altered by BMI is enriched for genes associated with cardiovascular and metabolic disease.

This study explored the effect of BMI on a comprehensive set of proteins in an MR framework. Previous studies have used observational epidemiology to explore the effect of obesity on the plasma proteome: one such study used mass spectrometry in two cohorts (19) and found higher BMI to be associated with seven out of 182 or 299 proteins (3.8% and 2.3% respectively), compared with 1,576 (39%) BMI-protein associations in the present study. An increase in Complement Factors I, B and H and an increase in CRP were observed (19). These findings were replicated in our current observational analysis using the SomaLogic platform, indicating that associations are detectable across different proteomic measurement platforms. The only association that did not replicate in the current study was the positive association with protein S100-A9. Although the MR analysis did not support some of these BMI-protein associations as being causal based on a P-value reference point, the strong association between the observational and MR estimates throughout the entire effect distribution suggests that disagreements between methods are likely an issue of power given current sample sizes.

For the proteins with stronger MR derived association evidence, it is important to explore (where possible) whether they have a potential role in disease. Our results suggest a strong positive effect of BMI on levels of leptin – a hormone released by white adipose tissue which suppresses appetite (38). The direction of effect we see currently agrees with estimates from previous cross-sectional and MR studies (17, 39). As higher levels of leptin are expected to reduce appetite and thus adiposity, this increase in circulating leptin seen with a higher BMI suggests that there is leptin receptor resistance (40). This observation illustrates the complexity encountered when interpreting MR estimates of causal effects and where, despite compelling association results, effect estimates can appear counter-intuitive. In this case, the positive relationship between BMI and an appetite regulating protein is a good illustration of the need to carefully explore the biology underpinning MR estimates before deploying them in more applied contexts (such as interventions). Knockout studies in mice have implicated higher leptin in arterial thrombosis (41) and there is observational evidence in humans that higher leptin can induce greater aggregation of platelets (cells involved in haemostasis) (42). In a larger population-based prospective observational study, leptin was found to be associated with higher risk of coronary events independent of BMI (43), whilst another observational study found leptin to be associated with higher risk of acute myocardial infarction (44).

Our results also suggest a strong positive effect of BMI on FABP4, an adipokine found primarily in adipocytes and macrophages (45). This association has been suggested in previous observational studies (46), but BMI-protein effect estimates from MR have been lacking. FABP4 has been implicated in cardiometabolic disease: a SNP which increases FABP4 was found to raise the odds of type II diabetes among adults (47), potentially through its contribution to higher insulin resistance (48). FABP has also been associated with higher risk of atherosclerosis among adults in an observational Chinese cohort study (49). A strong SHBG-lowering effect of higher BMI was also suggested here. The SHBG molecule is a glycoprotein which binds androgens and oestrogens and suppresses their activity (50); a reduction in SHBG is therefore expected to lead to higher levels of circulating sex hormones. The negative effect of BMI on SHBG seen here supports findings of previous cross-sectional studies among men (51) and post-menopausal women (52) and of a prospective study among post-menopausal women (53). When evaluating the role of SHBG in disease, one MR analysis suggests that a SNP which is associated with an increase in SHBG contributes to a decrease in risk of cardioembolic stroke (54), thereby suggesting that the SHBG-lowering effect of higher BMI observed in this study would increase this risk. Another study has also implicated lower SHBG levels in increasing type II diabetes risk (47). SHBG has also been implicated in cardiometabolic disease in observational studies. A study in over 3,000 men and women found that lower SHBG was associated with poorer cardiovascular health as assessed by a scale composed of cholesterol, glucose, blood pressure, BMI, diet, smoking, and physical activity (55). The exact mechanisms leading from decreased SBHG to ill-health is unclear, but may arise as a result of the increased bioavailability of testosterone and oestrogen (50).

Despite these possible protein involvements in cardiometabolic disease, it remains difficult to assess the contribution of individual proteins as they are not entirely independent and any pathological effects would likely be due to a global change in protein composition. There are not distinct groupings in the SomaLogic proteomics data as there often are with, for example, metabolomics data. We therefore additionally examined proteins grouped into clusters of similar features and compared BMI-protein estimates of each cluster with overall estimates and, for the cluster most affected by BMI, explored enrichment for genes related to disease. The cluster most altered by BMI (cluster 2) included most of the eight proteins with the strongest BMI effects from MR analyses, as well as various complement factors, chemokines, coagulation factors and IGFBPs, and was found to be enriched for genes related to cardiovascular disease, renal and metabolic diseases, and cancer. Enrichment was similar when comparing the proteins that had an observational association with BMI with all proteins included, with enrichment appearing greatest for renal and metabolic disease. Together, this suggests that changes in proteins identified here may mediate effects of obesity on cardiometabolic diseases; more focused investigations of these proteins are now needed.

This study has some limitations. Firstly, although INTERVAL is one of the largest existing cohorts to have untargeted proteomic data based on the SomaLogic platform, the sample size is still relatively modest and may have low power to detect some associations when using MR. Despite this, there was high agreement in the magnitude of effect estimates seen in observational and MR analyses which applied throughout the effect distribution. Participants with proteomic data were a small subset of the full INTERVAL cohort, however, this subset was randomly selected for proteomic assessment and comparisons between included and excluded participants revealed a similar profile in terms of age, sex, BMI, smoking, and alcohol use. Height and weight were self-reported which could introduce measurement error and bias; however, strong correlations are often reported between self-reported and measured BMI (58) and the validity of self-reported BMI here is supported by the strong positive association between the GRS for BMI (which was based largely on clinic-measured height and weight) and self-reported BMI in INTERVAL. Another limitation is the lack of availability of possible confounders for observational analyses. Although participants reported smoking and drinking habits, other potential confounders such as indicators of socio-economic position (which likely affect both BMI and protein traits related to cardiovascular disease processes (59, 60)) were not available. Residual confounding may help account for the much larger divergence between observed and expected P-values seen in observational versus MR models. The proteins examined are highly correlated and we therefore may not fully be describing changes in individual proteins. Evidence from case-control cohorts as well as functional and animal studies would help to isolate individual proteins that are altered and contribute to disease.

This study utilised SomaLogic to explore the relationship between BMI and plasma proteins in unprecendented scope and detail, in both an observational and MR framework. Here we provide evidence for a broad impact of higher adiposity on the human proteome. Causal evidence was strongest for BMI in relation to proteins involved in regulating appetite, sex hormones, and inflammation. Protein alterations were also found to be enriched for genes related to cardiovascular and metabolic disease. Altogether, these results help to focus attention onto new potential proteomic signatures of obesity-related disease. Further characterisation of the role of such proteomic profiles in disease using MR is warranted.

## Data Availability

Data cannot be publicly shared due to sensitive content. General enquiries can be sent to the INTERVAL team

https://www.intervalstudy.org.uk/more-information/

## Acknowledgments

Participants in the INTERVAL randomised controlled trial were recruited with the active collaboration of NHS Blood and Transplant England (www.nhsbt.nhs.uk), which has supported field work and other elements of the trial. DNA extraction and genotyping was co-funded by the National Institute for Health Research (NIHR), the NIHR BioResource (http://bioresource.nihr.ac.uk) and the NIHR [Cambridge Biomedical Research Centre at the Cambridge University Hospitals NHS Foundation Trust]*. The academic coordinating centre for INTERVAL was supported by core funding from: NIHR Blood and Transplant Research Unit in Donor Health and Genomics (NIHR BTRU-2014-10024), UK Medical Research Council (MR/L003120/1), British Heart Foundation (SP/09/002; RG/13/13/30194; RG/18/13/33946) and the NIHR [Cambridge Biomedical Research Centre at the Cambridge University Hospitals NHS Foundation Trust]. A complete list of the investigators and contributors to the INTERVAL trial is provided in this reference (1). The academic coordinating centre would like to thank blood donor centre staff and blood donors for participating in the INTERVAL trial.

This work was supported by Health Data Research UK, which is funded by the UK Medical Research Council, Engineering and Physical Sciences Research Council, Economic and Social Research Council, Department of Health and Social Care (England), Chief Scientist Office of the Scottish Government Health and Social Care Directorates, Health and Social Care Research and Development Division (Welsh Government), Public Health Agency (Northern Ireland), British Heart Foundation and Wellcome. This work was also supported by the Wellcome Trust grant number 206194.

*The views expressed are those of the authors and not necessarily those of the NHS, the NIHR or the Department of Health and Social Care.

