## Supplementary_figures for "Effects of adiposity on the human plasma proteome: Observational and Mendelian randomization estimates"

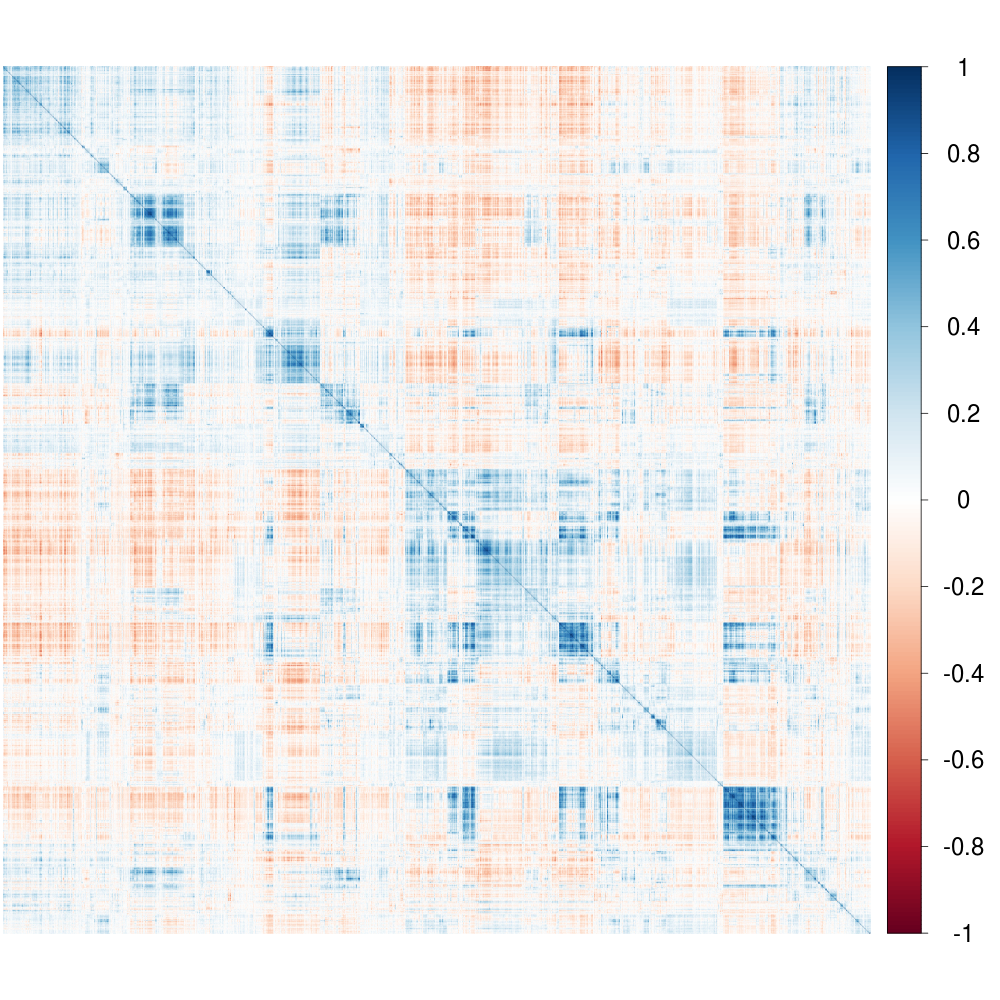

**Supplementary Fig 1**. Correlation matrix of all protein traits (4,034) where the colour corresponds to the correlation coefficient ranging from 1 (dark blue) to -1 (dark red).

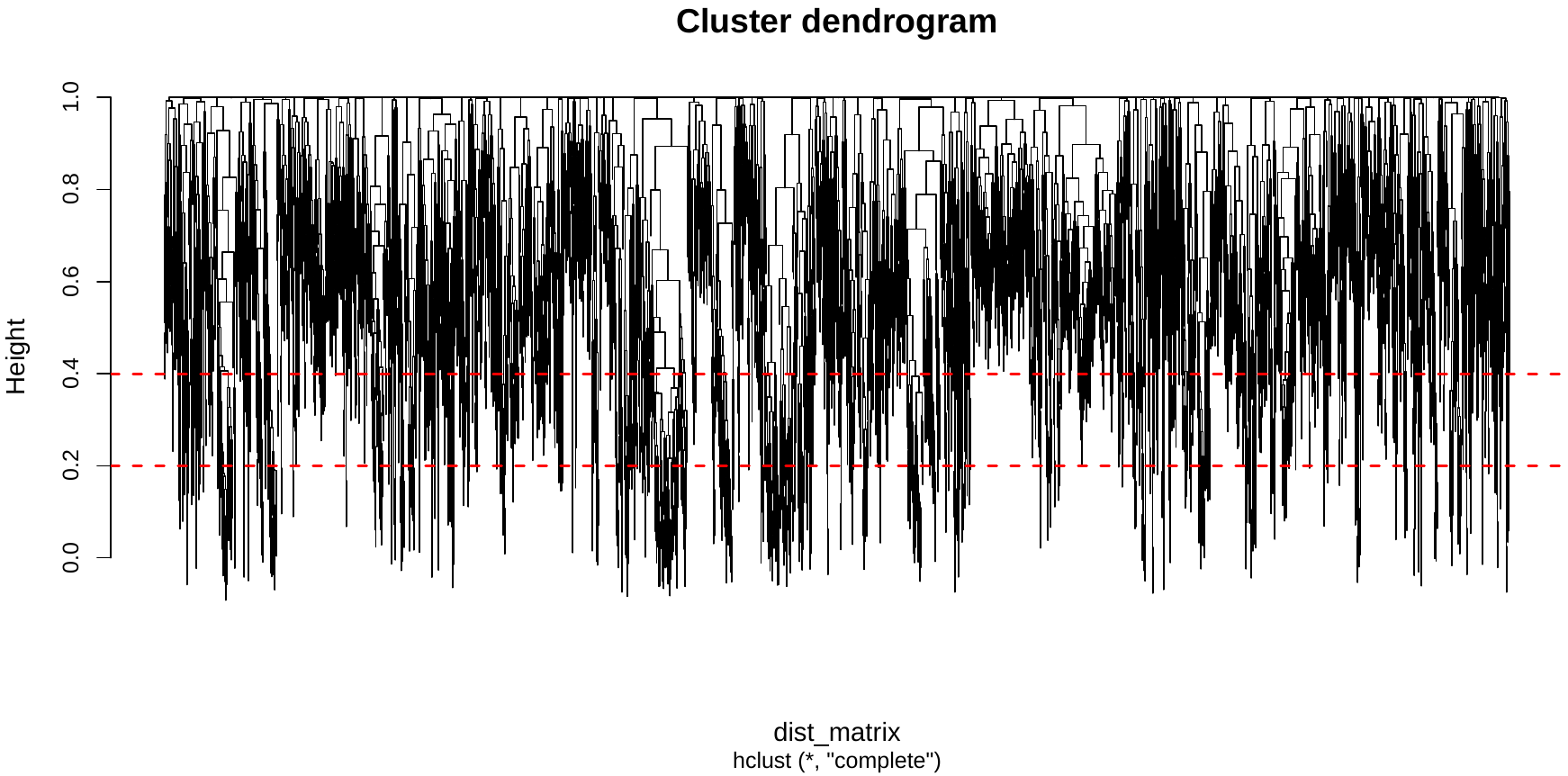

**Supplementary Fig 2.** Cluster dendrogram showing the hierarchal relationship between protein levels. Height is calculated as (1-correlation coefficient). At a height of 0.2 and 0.4 (red dashed lines), the number of independent proteins was 3,655 and 3,016 respectively.

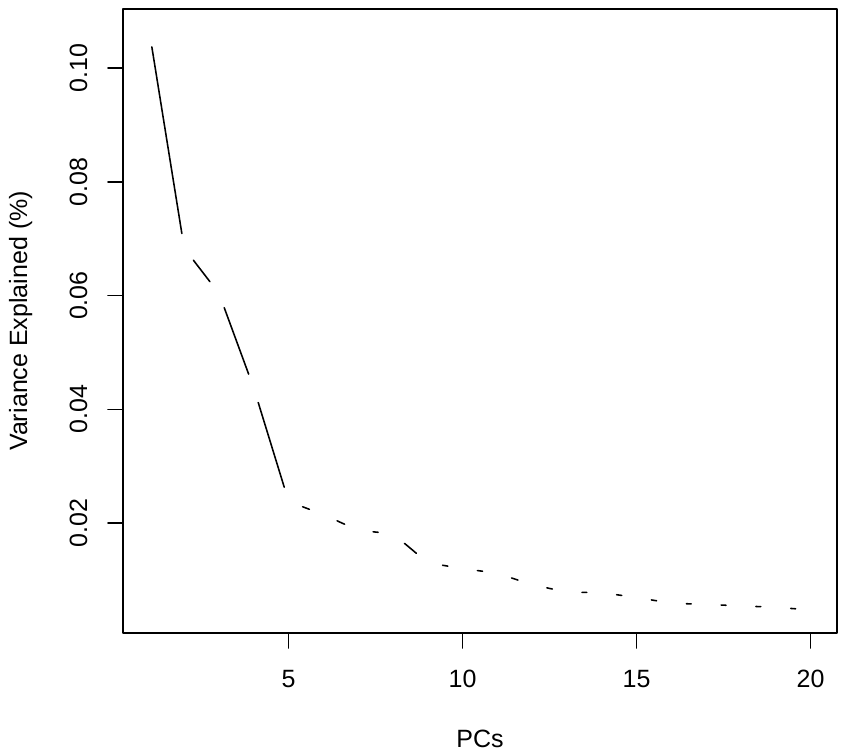

**Supplementary Fig 3**. Scree plot displaying the variance explained by each of the components in the principal component analysis (performed on the full protein dataset).

B

A

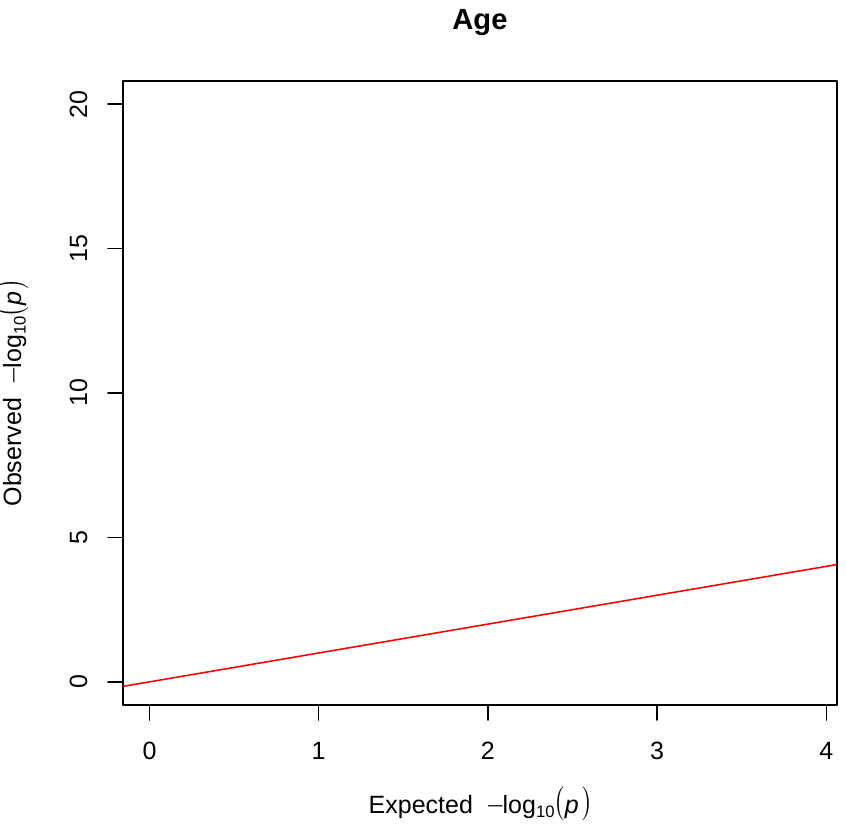

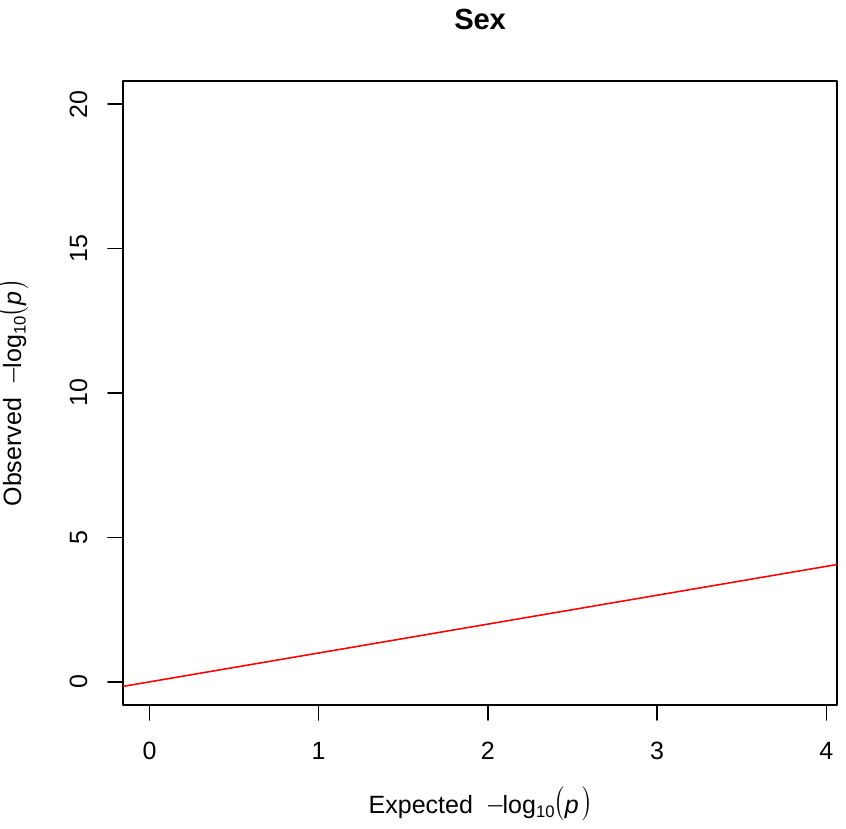

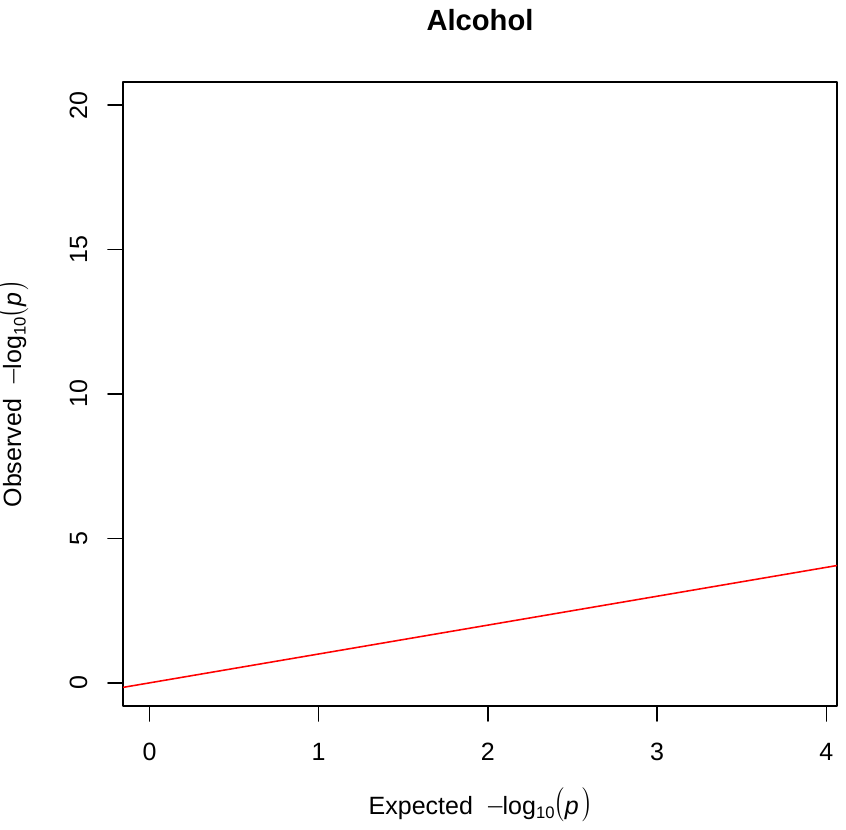

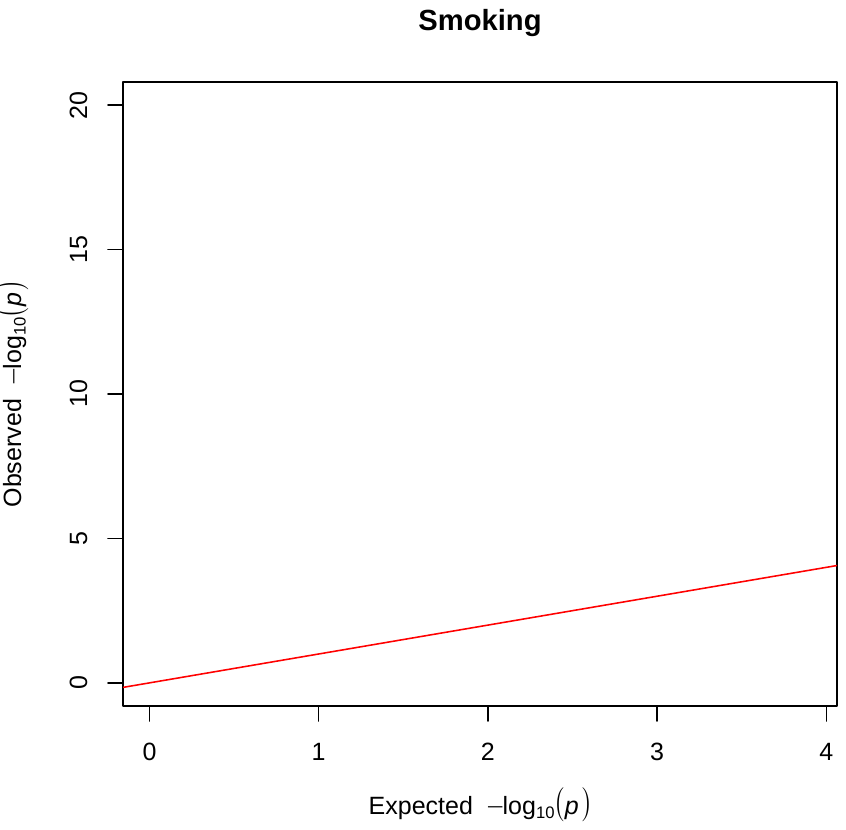

C
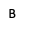

D

**Supplementary Fig 4.** Q-Q plots for the expected against observed -log_10_(p) values for the association of age (**A**), sex (**B**), smoking (**C**) and alcohol (**D**) with protein traits

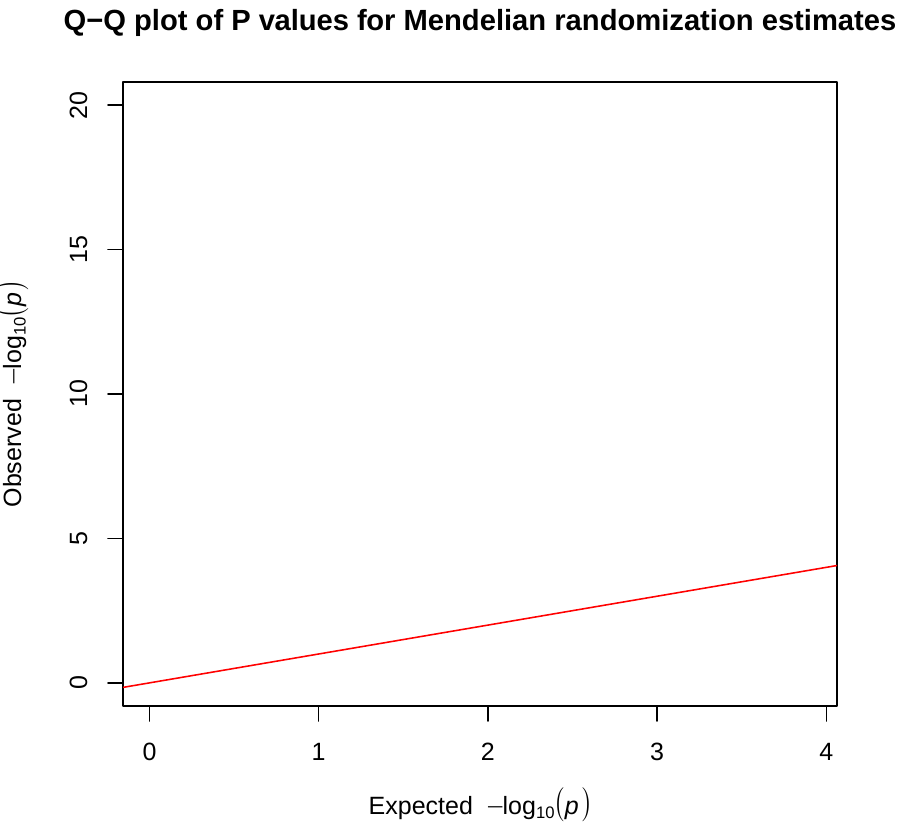

A

B

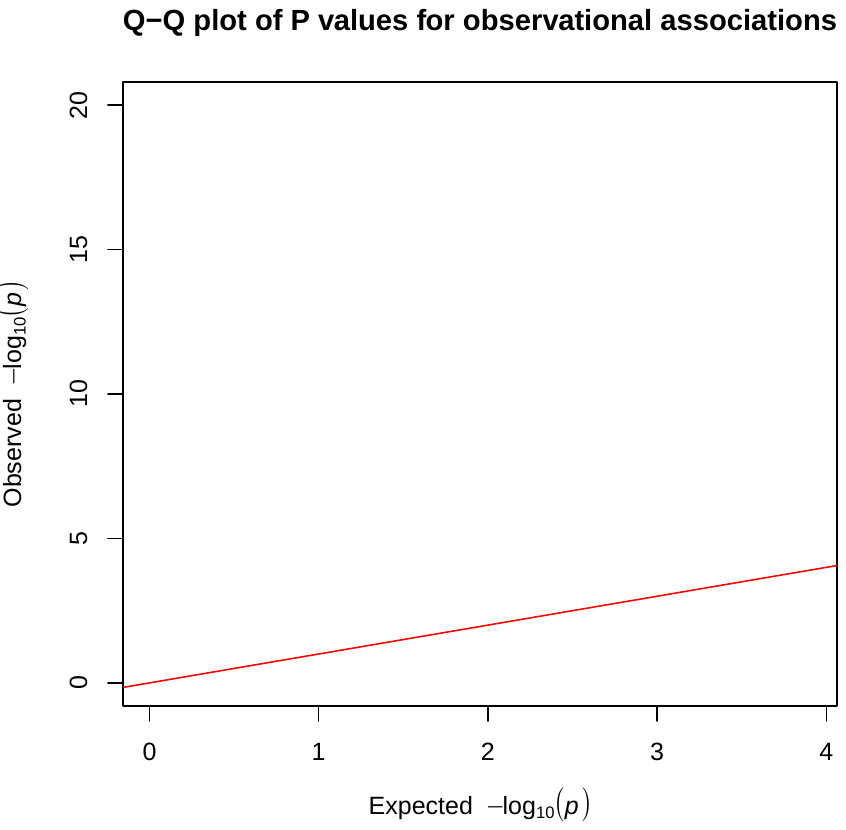

**Supplementary Fig 5. A**) Q-Q plot of expected against observed -log_10_(p) values for the unadjusted observational BMI-protein trait estimates **B**) Q-Q plot of expected against observed -log_10_(p) values for the Mendelian Randomization BMI-protein trait estimates.

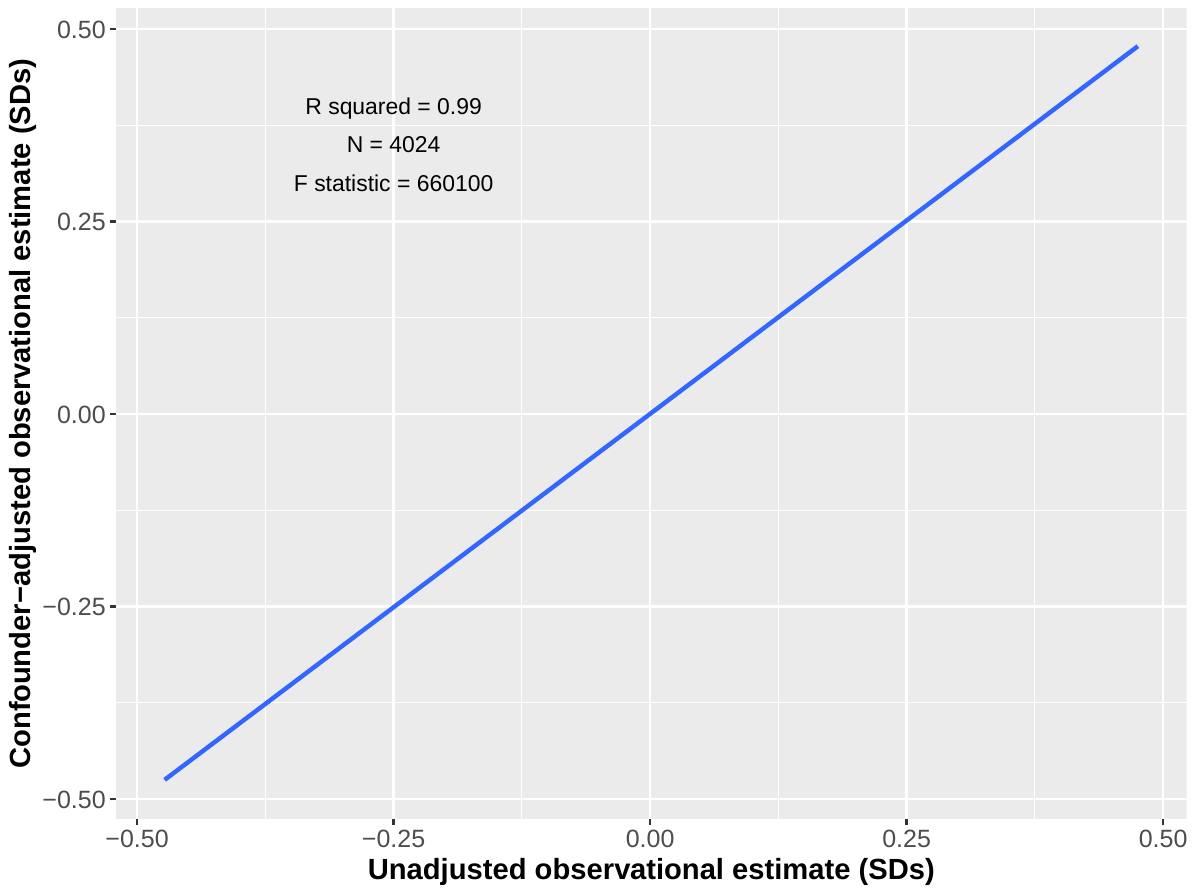

A

B

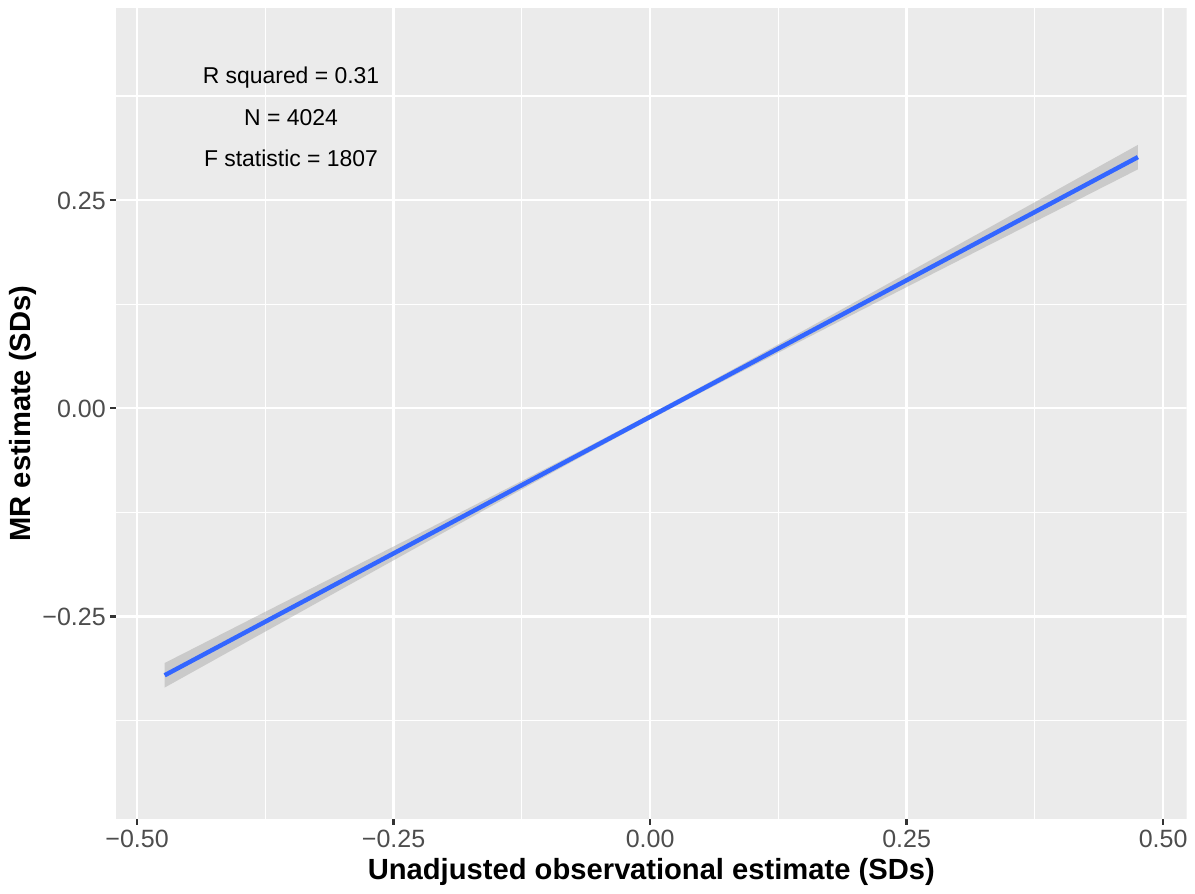

**Supplementary Fig 6. A)** Scatter plot of the unadjusted (age and sex adjusted) observational estimates and the confounder-adjusted observational estimates for BMI and protein traits with a regression line (blue), with the top eight MR BMI-associated proteins excluded. **6B**) Scatter plot of the unadjusted (age and sex adjusted) observational estimates and the MR estimates for BMI and protein traits with a regression line (blue), with the top eight MR BMI-associated proteins excluded.
